# Protective effects of testosterone replacement therapy on brain tumor outcomes: the Mayo Clinic Experience

**DOI:** 10.64898/2026.08.07.26359970

**Authors:** Manuel M. Bettencourt, Shreya Gandhi, Archis Bhandarkar, Asad Lone, Gelareh Zadeh, Sheila Mansouri

**Author notes:** Co-corresponding authors: **Dr. Sheila Mansouri, PhD** Associate Consultant I, Department of Neurologic Surgery, Assistant Professor of Neurosurgery, Associate Consultant, Mayo Clinic, Rochester, 200 First St. SW, Rochester, MN 55905, **Dr. Gelareh Zadeh, MD, PhD, FRCS(C)**, Chair, Department of Neurologic Surgery, Professor and Clinician Investigator of Neurosurgery, William J and Charles H Mayo Professor I, Chief Medical Officer, Mayo Clinic Platform, David C and Flora C Pratt Distinguished Chief Medical, Officer, Mayo Clinic, Rochester, 200 First St. SW, Rochester, MN 55905.

## Abstract

**Background:** Biological sex and endocrine signaling influence cancer biology, immune response, and therapeutic outcomes. Recent evidence suggests that testosterone signaling may exert brain-context-dependent protective effects in glioblastoma through the hypothalamic-pituitary-adrenal axis, reduced glucocorticoid-mediated immune suppression, and altered tumor-immune interactions. We assessed whether testosterone replacement therapy (TRT) exposure was associated with survival in solid tumor central nervous system (CNS) metastases and glioblastoma (GBM, IDH-wildtype, WHO grade 4), settings in which post-diagnosis survival and TRT timing can be clinically defined.

**Methods:** We performed a retrospective Mayo Clinic cohort study of adult patients with molecularly confirmed glioblastoma and solid tumor CNS metastases confirmed from neuroimaging reports using large language model-assisted adjudication. TRT exposure was defined by testosterone-specific prescription evidence within prespecified peri-diagnostic windows. Overall survival was evaluated using propensity score-matched Cox models, 24-month administratively censored Cox models, time-dependent Cox sensitivity analyses, and 24-month restricted mean survival time.

**Results:** In the pooled solid tumor CNS metastasis cohort, TRT exposure was associated with improved overall survival after propensity score matching (HR 0.80, 95% CI 0.65–0.98, p=0.029) and a 3.22-month improvement in 24-month restricted mean survival time. In glioblastoma, TRT exposure was similarly associated with improved overall survival after propensity score matching (HR 0.56, 95% CI 0.38–0.82, p=0.003) and a 5.81-month improvement in 24-month restricted mean survival time.

**Conclusions:** TRT exposure was associated with improved survival in CNS metastases and glioblastoma. These hypothesis-generating findings support prospective studies incorporating TRT timing, hormone levels, corticosteroid exposure, immune correlates, and tumor-specific stratification.

**Key points:**

- TRT exposure is associated with improved survival in aggressive brain tumors.
- Survival associations persist after propensity score matching.
- Prospective studies should define TRT timing, hormone levels, and immune effects.

**Importance of Study:** This study evaluates testosterone replacement therapy (TRT) in aggressive intracranial tumors with clinically interpretable survival timelines. Building on recent evidence that androgen signaling may have protective brain-specific effects in glioblastoma, we extend this question to CNS metastases. In a multi-site Mayo Clinic cohort, TRT exposure was associated with improved survival in CNS metastases and glioblastoma, supporting androgen biology as an underrecognized factor in neuro-oncology outcomes. These findings provide rationale for prospective clinical trials incorporating TRT timing, hormone levels, corticosteroid exposure, immune profiling, and tumor-specific stratification.

## Introduction

Age and sex are major biological determinants of incidence, progression, therapeutic response, and survival in most tumors^1–4^. Although these factors are often analyzed independently, they are biologically connected through endocrine, immune, metabolic, and epigenetic pathways^5,6^. While age is associated with hormone levels, sex differences in cancer are also shaped by sex chromosome dosage, X-chromosome inactivation or escape, mosaic Y-chromosome loss, and tissue-specific responses to sex hormones, including androgen and estrogen^7,8^. These factors influence both tissue intrinsic programs and host immune responses. The central nervous system (CNS) provides a unique context in which to study these interactions. Unlike most peripheral tissues, the brain contains specialized immune, vascular, metabolic, and neuroendocrine components, including brain resident microglia, blood-brain barrier-associated endothelial cells, and direct coupling to hypothalamic-pituitary-adrenal (HPA) axis signaling^9,10^. The HPA axis coordinates systemic stress responses through glucocorticoid release, which can suppress antitumor immunity, alter myeloid cell states, and influence tumor-host interactions within the brain microenvironment^11–13^. Sex-dependent developmental programs also influence neural progenitor cell states and shape tumor biology in brain tumors^14^.

Glioblastoma (GBM, IDH-wildtype, WHO grade 4) is a prominent example of the clinical relevance of sex differences in neuro-oncology. Glioblastoma demonstrates both higher incidence rates and worse outcomes in males relative to females^15,16^. Prior studies have identified sex-associated differences in glioblastoma biology^17^, including variations in tumor growth kinetics, immune suppression, transcriptional programs, myeloid-derived suppressor cell biology^18^, T cell function^19^, and microglial tumor-suppressive mechanisms, including a role for junctional adhesion molecule A as a female-biased microglial tumor suppressor^20–24^. Altogether, these findings suggest that biological sex influences glioblastoma biology through both tumor-intrinsic and immune mechanisms.

Androgen signaling may be particularly relevant in this context because androgens regulate brain development, immune function, and neuroendocrine signaling, and testosterone can be locally converted to estrogens through aromatase^25–30^. As a result, the effects of testosterone or androgen receptor (AR) signaling within the CNS may not be consistent with their effects in systemic malignancies. Studies in prostate, bladder, and colon cancer have demonstrated that androgen deprivation or AR blockade can enhance anti-tumor immunity^31,32^. Similarly, *in vitro* glioblastoma studies have suggested that testosterone promotes glioma cell proliferation, migration, invasion, and radiation resistance^33,34^. Additional evidence further links AR signaling to glioblastoma growth and therapeutic resistance. AR is amplified or overexpressed in a subset of glioblastoma^35^. EGFR/EGFRvIII signaling can also promote ligand-independent AR activation^36^. Consistent with these findings, AR inhibition or degradation reduces glioblastoma growth in preclinical models and prolongs survival in xenograft studies^35,37,38^. These observations have contributed to the conventional view that androgen signaling is immunosuppressive and tumor-promoting in cancer.

A recent landmark study by Lee et al. challenged this paradigm^39^, demonstrating that although testosterone enhanced the proliferation of glioma cells *in vitro,* in murine glioma castration model androgen deprivation paradoxically accelerated intracranial tumor growth, whereas testosterone supplementation delayed tumor progression and improved survival. Mechanistically, androgen loss activated the HPA axis, increased systemic glucocorticoids, impaired T cell function, and promoted a myeloid-dependent immunosuppressive microenvironment. These findings support a model in which testosterone exerts context-dependent, brain-specific protective neuroimmune effects. However, whether these observations extend beyond glioblastoma remains unknown. Furthermore, brain tumors are biologically heterogeneous, and the relationship between testosterone replacement therapy (TRT) and outcome may differ in brain metastases. In metastatic disease, these effects may be further influenced by primary cancer type and interactions with systemic therapies, including AR-directed agents and immune checkpoint inhibitors (ICIs).

Here, we evaluate the association between TRT exposure and survival in glioblastoma and solid-tumor CNS metastases using a multi-site Mayo Clinic cohort accessed through the Mayo Clinic Platform. These disease contexts were selected because their post-diagnosis survival trajectories allow clinically meaningful definition of TRT exposure windows and survival endpoints. We assessed whether TRT-associated survival differences persisted after adjustment for clinically relevant covariates and performed sensitivity analyses to address treatment timing. In patients with CNS metastases, we further examined whether TRT exposure was associated with survival after metastasis diagnosis and explored potential interactions with immune checkpoint inhibitor therapy. Together, these analyses extend recent evidence that androgen biology may influence intracranial tumor outcomes through mechanisms distinct from those observed in systemic cancers.

## Materials and Methods

### Study design and cohorts

We performed retrospective cohort analyses of male patients evaluated at Mayo Clinic using structured and semi-structured electronic health record (EHR) data in the Mayo Clinic Platform. Two analytic cohorts were defined: a CNS metastasis cohort and a glioblastoma cohort (GBM, IDH-wildtype, WHO grade 4). Cohort characteristics are summarized in **Supplementary Table 1**.

For the CNS metastasis cohort, eligible patients were > 18 years of age at diagnosis, male, had a documented primary malignancy, known vital status, available survival follow-up, and strict evidence of CNS metastatic disease. CNS metastasis was confirmed from dedicated neuroimaging reports adjudicated using a large language model (LLM) and prespecified text rules. Reports were considered confirmatory only when they contained affirmative evidence of brain, leptomeningeal, spinal CNS, or other intracranial metastatic disease. Reports were excluded as confirmatory when the LLM identified negated disease, extracranial-only metastatic disease, nonbrain metastases only, or insufficient evidence of CNS involvement. Residual or persistent CNS disease was counted only when explicitly anchored to metastatic disease, metastases, leptomeningeal disease, leptomeningeal carcinomatosis, or known CNS metastatic disease. Ambiguous residual enhancement, unanchored residual tumor, treatment effect, necrosis favored over metastasis, or concern for a primary CNS tumor was not considered confirmatory. Hard-negative phrases, including “no evidence of metastasis,” were evaluated using LLM interpretation fields rather than the full report body to avoid excluding reports with true positive metastatic language. Patient-level confirmation required at least one strict positive report. No broad manual chart review was performed beyond targeted review of cases affected by hard-negative language logic.

The glioblastoma cohort was derived from the cancer registry and EHR diagnosis fields, with the cancer registry glioblastoma diagnosis date used as the index date. Eligible patients were ≥18 years of age, male, had registry or EHR evidence of glioblastoma with IDH status determined from cancer registry data, known vital status, available survival follow-up, and MGMT promoter methylation status classified as methylated or unmethylated. MGMT status was derived from registry molecular fields; absent, not present, or unmethylated values were classified as unmethylated, whereas present, low-level, high-level, or level-unspecified methylation values were classified as methylated.

### Definition of TRT exposure

TRT exposure was derived from testosterone-specific medication evidence. For the CNS metastasis analysis, TRT exposure was defined as any qualifying TRT evidence overlapping the interval from 3 months before through 12 months after CNS metastasis diagnosis. For the glioblastoma analysis, the primary TRT exposure flag included TRT during the temozolomide (TMZ) treatment window or TRT within the registry-diagnosis-centered window from 3 months before through 12 months after glioblastoma diagnosis. Broad route or formulation terms, such as injection, syringe, vial, infusion, or pump, were not considered sufficient unless directly anchored to testosterone medication evidence.

### Definition of outcome

The primary outcome in both cohorts was overall survival. In the CNS metastasis cohort, survival was measured from CNS metastasis diagnosis to death or last follow-up. In the glioblastoma cohort, survival was measured from glioblastoma diagnosis to death or last follow-up. Patients without documented death were censored at last follow-up. For both cohorts, we evaluated overall survival over full available follow-up and also created a 24-month administratively censored endpoint, in which follow-up was truncated at 24 months and only deaths occurring within 24 months were counted as events. Restricted mean survival time was estimated through 24 months in both cohorts. Kaplan-Meier curves were used for descriptive visualization: the CNS metastasis cohort was displayed using the 24-month censored analysis, whereas the glioblastoma cohort was displayed as overall survival with the x-axis truncated at 36 months.

### Propensity score matching

For the pooled CNS metastasis analysis, the propensity-score model included age at CNS metastasis diagnosis, primary cancer group, overall extracranial metastatic disease, post-diagnosis radiation therapy, stereotactic radiosurgery, craniotomy or resection, and any local CNS-directed therapy. For the glioblastoma analysis, the propensity score matching model (PSM) included age at diagnosis, MGMT methylation status, TMZ exposure, dexamethasone exposure, and radiation procedure evidence.

Propensity scores were estimated using logistic regression with TRT exposure as the dependent variable. Both analyses used full matching without replacement as implemented in the ‘MatchIt’ R package. For the pooled CNS metastasis analysis, full matching used a standardized propensity-score caliper of 0.2. For the glioblastoma analysis, full matching used a standardized PSM caliper of 0.005. Covariate balance was assessed using standardized mean differences before and after matching. Absolute standardized mean differences below 0.10 were considered acceptable. Matched analyses used matching weights, and Cox models used robust standard errors.

### Survival analyses

The primary model in each cohort was a weighted Cox proportional hazards model fit in the matched cohort, estimating the association between TRT exposure and overall survival. Supportive analyses included 24-month administratively censored Cox models and 24-month restricted mean survival time (RMST) analyses. RMST was used to estimate the difference in mean survival time restricted to the first 24 months. For the pooled CNS metastasis cohort, an additional time-dependent Cox sensitivity analysis was performed to address potential immortal-time bias related to TRT initiation after diagnosis. Patients with TRT before or at diagnosis were considered exposed from time zero. Patients starting TRT after diagnosis contributed unexposed person-time until the first TRT date and exposed person-time thereafter. Patients without TRT contributed only unexposed person-time. The primary time-dependent sensitivity analysis was covariate-adjusted and administratively censored at 24 months.

### Statistical analysis

Continuous variables were summarized using medians and interquartile ranges. Categorical variables were summarized using counts and percentages. Cox model results are reported as hazard ratios with 95% confidence intervals. Restricted mean survival time (RMST) results are reported as restricted mean survival time differences in months with 95% confidence intervals. Kaplan-Meier plots included median survival estimates, and descriptive log-rank p-values; primary inference was based on matched Cox and RMST analyses rather than unweighted log-rank tests. Analyses were performed in R.

## Results

### Cohort characteristics

The clinical and treatment characteristics of the CNS metastasis and matched glioblastoma cohorts are summarized in **Supplementary Table 1.** The pooled solid tumor CNS metastasis cohort included 5,287 male patients, of whom 91 were TRT-exposed and 5,196 were not TRT-exposed. Overall, 4,293 deaths occurred, and the median survival from CNS metastasis diagnosis was 7.19 months. Lung cancer was the most common primary tumor type in both groups, representing 3,000 No-TRT patients and 43 TRT-exposed patients. Other primary tumor types included melanoma, renal, colorectal, gastrointestinal/hepatobiliary/pancreatic, urothelial, and sarcoma. TRT exposure varied by primary tumor type and was most frequent among patients with renal cancer, where 17 of 555 patients were TRT-exposed, followed by melanoma with 21 of 976 patients, and urothelial cancer with 2 of 105 patients.

Patients in the CNS metastasis cohort were similar in age at CNS metastasis diagnosis, with a median age of 66.0 years in the No-TRT group and 67.7 years in the TRT group. TRT-exposed patients had a median first TRT order 27 days before CNS metastasis diagnosis. Extracranial metastatic disease was documented in 61.5% of TRT-exposed patients and 50.6% of No-TRT patients. Radiation exposure was similar in both groups, occurring in 69.2% of TRT-exposed patients and 68.6% of No-TRT patients. Stereotactic radiosurgery was documented in 39.6% of TRT-exposed patients and 36.2% of No-TRT patients, while resection or craniotomy was documented in 17.6% and 14.3%, respectively. Any form of local CNS metastasis-directed therapy was documented in 71.4% of TRT-exposed patients and 72.6% of No-TRT patients. Death occurred in 78.0% of TRT-exposed patients and 81.3% of No-TRT patients.

The glioblastoma cohort included 598 male patients, comprising 11 TRT-exposed and 587 No-TRT patients. Median age at glioblastoma diagnosis was 68.2 years in the TRT group and 64.6 years in the No-TRT group. In regard to MGMT promoter methylation status, methylation was present in 36.4% of TRT-exposed patients and 33.7% of No-TRT patients. Treatment exposures were also broadly similar: TMZ exposure occurred in 72.7% of TRT-exposed patients and 79.0% of No-TRT patients, dexamethasone exposure in 72.7% and 87.9%, radiation procedures in 81.8% and 69.7%, and surgery in 90.9.0% and 85.9%, respectively. Deaths occurred in 63.6% of TRT-exposed patients compared with 86.9% of No-TRT patients. Median survival was 12.6 months in the TRT group and 13.2 months in the No-TRT group.

Performance status was summarized descriptively across cohorts using available KPS and ECOG data. In the CNS metastasis cohort, ECOG was available for most patients, with ECOG 0–1 documented in 43.4% of No-TRT and 51.6% of TRT-exposed patients; ECOG was missing in 41.6% and 36.3%, respectively. KPS was less frequently available, with missing values in 87.3% of No-TRT and 85.7% of TRT-exposed patients. In the glioblastoma cohort, ECOG was available in most patients, with missing values in 28.8% of No-TRT and 27.3% of TRT-exposed patients, while KPS was missing in 52.5% and 36.4%, respectively. Because performance status was captured incompletely and across different scoring systems, KPS and ECOG were not included in the propensity score models.

### TRT exposure is associated with improved survival post-CNS metastases diagnosis

We next evaluated whether TRT exposure was associated with overall survival after diagnosis of solid tumor CNS metastases. PSM was used to balance TRT and no-TRT patients across clinically relevant baseline and treatment variables, including age at CNS metastasis diagnosis, primary tumor type, extracranial metastatic disease, radiation, stereotactic radiosurgery, craniotomy, and other local CNS-directed therapies. Covariate balance improved after matching, with standardized mean differences less than 0.1 across matched variables (**Figure 1A**).

**Figure 1.**
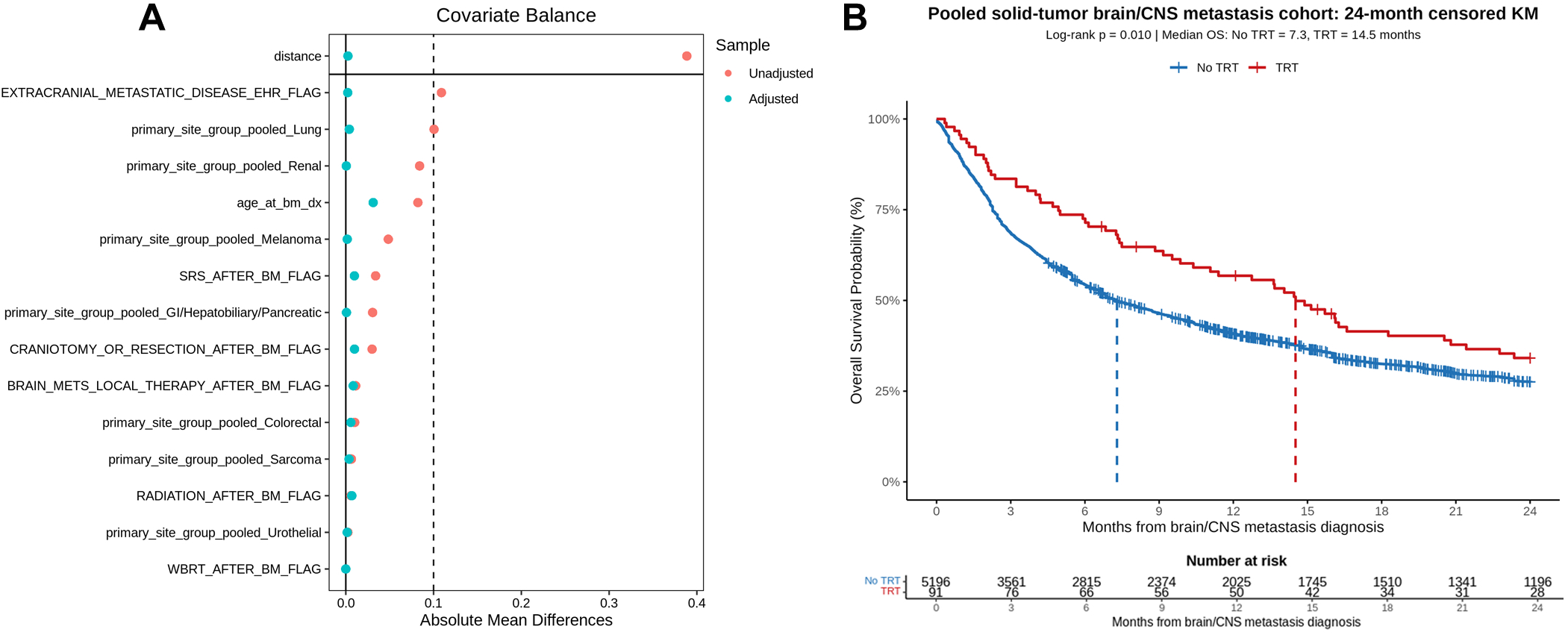
TRT exposure is associated with improved survival after solid tumor CNS metastasis diagnosis. **A**, Covariate balance before and after PSM in the pooled solid tumor CNS metastasis cohort. Absolute standardized mean differences are shown for variables included in the propensity score model; the dashed vertical line indicates the 0.10 balance threshold. **B**, Twenty-four-month administratively censored Kaplan-Meier curves comparing overall survival after CNS metastasis diagnosis in TRT-exposed and unexposed patients. Number-at-risk tables are shown below the curves. HR, hazard ratio; CI, confidence interval.

In the primary PSM Cox analysis, TRT exposure was associated with a significantly lower hazard of death compared with no TRT exposure (HR 0.80, 95% CI 0.65-0.98, *p* = 0.029). This association remained significant in a 24-month administratively censored Cox model, which focused inference on the early post-metastasis survival interval and reduced the influence of long-term survival outliers (HR 0.74, 95% CI 0.58-0.93, *p* = 0.012).

Because TRT exposure could occur before or after CNS metastasis diagnosis, we performed a 24-month time-dependent Cox sensitivity analysis to account for treatment timing and reduce potential immortal-time bias. In this analysis, patients contributed unexposed person-time until the first documented TRT order and exposed person-time thereafter. The effect pattern remained consistent with the primary analysis, although it did not reach statistical significance (HR 0.80, 95% CI 0.61-1.05, *p* = 0.110).

Kaplan-Meier analysis of the matched cohort demonstrated longer survival among TRT-exposed patients compared with unexposed patients after CNS metastasis diagnosis (**Figure 1B**). To quantify the absolute survival difference, we estimated restricted mean survival time through 24 months. TRT exposure was associated with a 3.22-month improvement in 24-month restricted mean survival time compared with no TRT exposure (95% CI 1.35-5.09 months, *p* < 0.001). These analyses show that TRT exposure was associated with improved survival after CNS metastasis diagnosis, with consistent directionality across PSM Cox, 24-month censored Cox, time-dependent Cox, Kaplan-Meier, and restricted mean survival time analyses.

### Immune checkpoint blockade does not significantly modify the TRT survival association in CNS metastases

Given the known effects of androgen signaling on systemic immune regulation, we next evaluated whether the association between TRT exposure and survival differed by immune checkpoint inhibitor (ICI) exposure status in the CNS metastasis cohort. In main-effects Cox models, adjusted for the prespecified CNS metastasis covariates, both TRT exposure and ICI exposure were independently associated with improved overall survival. TRT exposure was associated with lower mortality risk (HR 0.78, 95% CI 0.63-0.97, *p* = 0.025), and ICI exposure was similarly associated with improved survival (HR 0.84, 95% CI 0.78-0.90, *p* < 0.001).

Among ICI-exposed patients, Kaplan-Meier analysis showed numerically longer median overall survival in TRT-exposed patients compared with unexposed patients, although this difference was not statistically significant by log-rank testing (16.1 vs 10.7 months, *p* = 0.354; **Fig. 2**). Specifically, interaction testing demonstrated no significant TRT × ICI interaction in the overall Cox model (HR 1.09, 95% CI 0.72-1.66, *p* = 0.684), indicating that the association between TRT exposure and survival was not significantly modified by ICI exposure. Similar findings were observed in the 24-month interaction model, where the TRT × ICI interaction remained non-significant (HR 1.04, 95% CI 0.63-1.72, *p* = 0.863).

**Figure 2.**
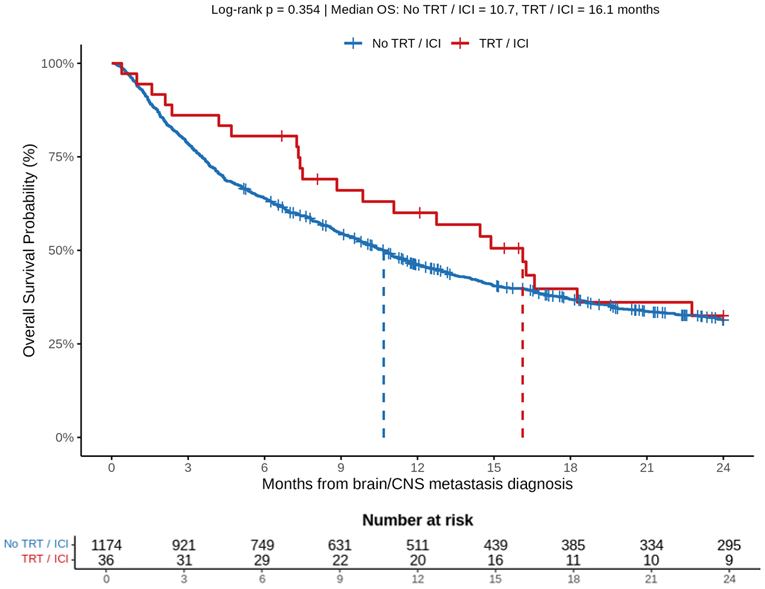
ICI exposure does not significantly modify the TRT-survival association in CNS metastases. Kaplan-Meier curves of overall survival among ICI-exposed patients stratified by TRT exposure, with number-at-risk tables shown below.

In stratified interaction models, TRT exposure among ICI-negative patients was associated with a directionally favorable but borderline non-significant survival estimate in the overall model (HR 0.76, 95% CI 0.56-1.01, *p* = 0.061), and reached significance in the 24-month model (HR 0.71, 95% CI 0.51-0.99, p = 0.045). ICI exposure among non-TRT patients was consistently associated with improved survival in both the overall model (HR 0.84, 95% CI 0.77-0.90, *p* < 0.001) and the 24-month model (HR 0.83, 95% CI 0.76-0.90, *p* < 0.001) (**Table 1**). These findings suggest that TRT and ICI exposures were each associated with improved survival in CNS metastases, but ICI exposure did not significantly enhance or attenuate the survival association of TRT.

**Table 1.** TRT and ICI associations with overall survival in CNS metastases. Cox proportional hazards models evaluating the independent and interactive associations of TRT and ICI exposure with overall survival in the CNS metastasis cohort. Hazard ratios are adjusted for prespecified CNS metastasis covariates. Interaction terms test whether the association between TRT exposure and survival differs by ICI exposure status. HR, hazard ratio; CI, confidence interval; CNS, central nervous system; TRT, testosterone replacement therapy; ICI, immune checkpoint inhibitor.

| Section | Model | Analysis | HR (95% CI) | P value |
| --- | --- | --- | --- | --- |
| Main effects model | Main-effects Cox | TRT main effect | **0.78 (0.63-0.97)** | **0.025** |
| Main effects model | Main-effects Cox | ICI main effect | **0.84 (0.78-0.90)** | **<0.001** |
| Overall interaction model | Interaction Cox | TRT effect among ICI-negative patients | 0.76 (0.56-1.01) | 0.061 |
| Overall interaction model | Interaction Cox | ICI effect among non-TRT patients | **0.84 (0.77-0.90)** | **<0.001** |
| Overall interaction model | Interaction Cox | TRT × ICI interaction | 1.09 (0.72-1.66) | 0.684 |
| 24-month interaction model | 24-month interaction Cox | 24-month TRT effect among ICI-negative patients | **0.71 (0.51-0.99)** | **0.045** |
| 24-month interaction model | 24-month interaction Cox | 24-month ICI effect among non-TRT patients | **0.83 (0.76-0.90)** | **<0.001** |
| 24-month interaction model | 24-month interaction Cox | 24-month TRT × ICI interaction | 1.04 (0.63-1.72) | 0.863 |

### TRT exposure is associated with improved survival in glioblastoma patients

We next evaluated whether TRT exposure was associated with survival in male patients with glioblastoma. Full PSM was used to balance TRT-exposed and unexposed patients across clinically relevant covariates, including age at glioblastoma diagnosis, MGMT promoter methylation status, TMZ exposure, dexamethasone exposure, and radiation procedure. Covariate balance improved after matching, with adjusted standardized mean differences less than 0.1 across matched variables (**Figure 3A**).

**Figure 3.**
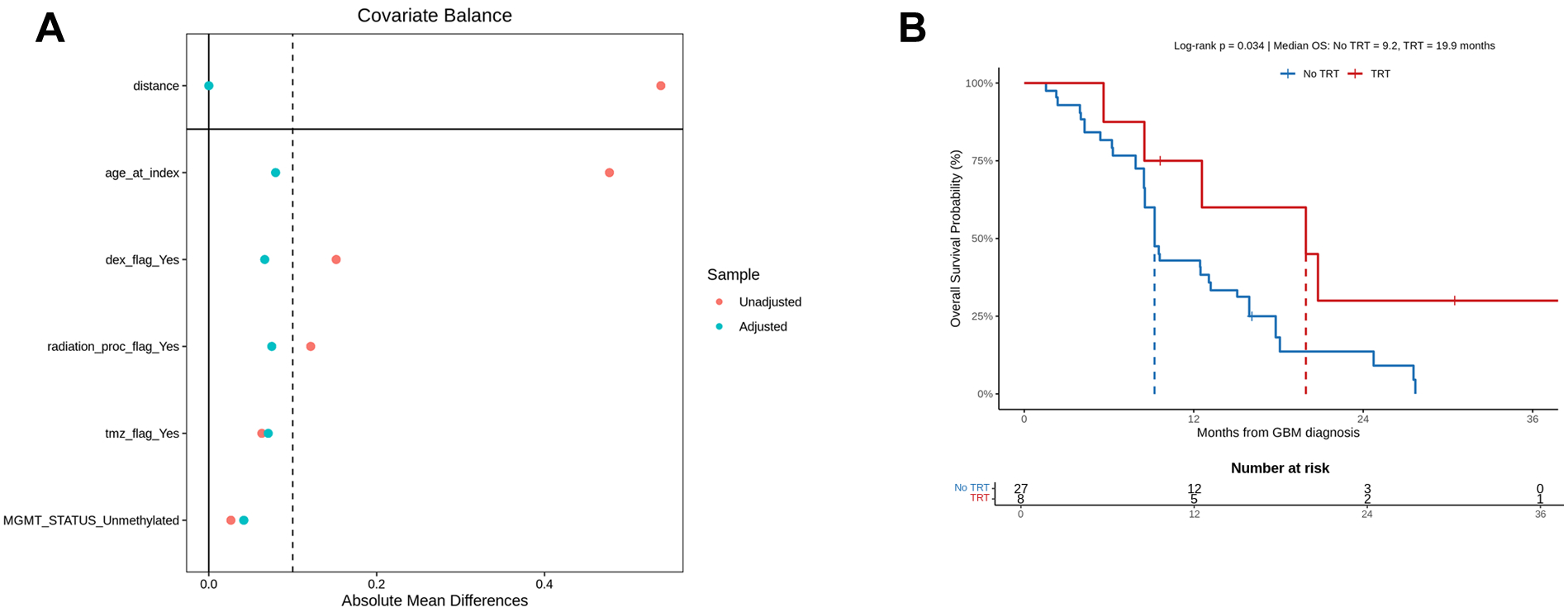
TRT exposure is associated with improved survival in male patients with glioblastoma. **A**, Covariate balance before and after full propensity score matching in the glioblastoma cohort. Absolute standardized mean differences are shown for variables included in the propensity score model; the dashed vertical line indicates the 0.10 balance threshold. **B**, Kaplan-Meier curves comparing overall survival from glioblastoma diagnosis in TRT-exposed and unexposed patients in the matched cohort. Number-at-risk tables are shown below the curves. MGMT, O6-methylguanine-DNA methyltransferase.

The matched glioblastoma cohort included 35 patients, comprising 8 TRT-exposed and 27 unexposed patients. Baseline and treatment characteristics were broadly similar between groups after matching. Median age at glioblastoma diagnosis was 64.2 years in TRT-exposed patients and 61.2 years in unexposed patients. MGMT promoter methylation was present in 37.5% of TRT-exposed patients and 33.3% of unexposed patients. TMZ exposure occurred in 75.0% of TRT-exposed patients and 70.4% of unexposed patients, dexamethasone exposure in 87.5% and 81.5%, radiation procedures in 75.0% and 66.7%, and surgery in 100.0% and 96.3%, respectively.

In the primary PSM Cox analysis, TRT exposure was associated with a significantly lower hazard of death compared with no TRT exposure (HR 0.38, 95% CI 0.14-0.98, *p* = 0.046). This association remained significant in the 24-month administratively censored Cox analysis, which focused inference on the clinically relevant early survival interval for glioblastoma (HR 0.48, 95% CI 0.24-0.99, *p* = 0.048).

Kaplan-Meier analysis of the matched cohort showed longer overall survival among TRT-exposed patients compared with unexposed patients after glioblastoma diagnosis (log-rank *p* = 0.034; **Figure 3B**). Median overall survival was 19.9 months in TRT-exposed patients compared with 9.2 months in unexposed patients. To quantify the absolute survival difference, we estimated restricted mean survival time through 24 months. TRT exposure was associated with a 5.81-month improvement in 24-month restricted mean survival time compared with no TRT exposure (95% CI 0.31-11.30 months, *p* = 0.038). These results demonstrate that TRT exposure was associated with improved survival in matched male patients with glioblastoma.

## Discussion

In this retrospective multi-site Mayo Clinic cohort study, TRT exposure was associated with improved overall survival in male patients with solid tumor CNS metastases and glioblastoma. In CNS metastases, this association remained consistent after PSM and was supported by 24-month censored Cox and restricted mean survival time analyses. A consistent pattern was observed after time-dependent sensitivity testing. In glioblastoma, TRT exposure was also associated with longer survival after full PSM and a 5.81-month improvement in 24-month restricted mean survival time. These findings support the hypothesis that androgen biology may influence outcomes in aggressive intracranial tumors with clinically definable survival, follow-up, and treatment windows.

Our findings in glioblastoma are consistent with recent experimental evidence showing that androgen loss may accelerate intracranial tumor growth through HPA axis activation, glucocorticoid-mediated immune suppression, impaired T-cell function, and myeloid remodeling^39^. The CNS metastasis findings extend this observation beyond glioblastoma, suggesting that even tumors originating outside the CNS may be influenced by the intracranial immune and endocrine environment after metastatic seeding.

We also examined whether ICI exposure modified the TRT-survival association in CNS metastases. TRT and ICI exposure were both independently associated with improved overall survival in adjusted models. However, formal TRT × ICI interaction testing showed no significance, suggesting that the TRT-associated survival advantage was not significantly enhanced or attenuated by ICI exposure. Larger studies with tumor-specific stratification and immune profiling should be performed to determine whether androgen signaling modifies immunotherapy response in CNS metastases.

This study focused on glioblastoma and solid tumor CNS metastases because these diseases have short and clinically interpretable survival timelines, allowing TRT exposure to be defined around diagnosis and early treatment. Other intracranial tumors, such as meningiomas and gliomas, were not included because longer natural histories, lower event rates, and difficulty defining TRT timing relative to diagnosis or treatment would limit data interpretation. This restriction improves the interpretability of TRT exposure timing and aligns the analysis with the type of defined treatment window needed for prospective clinical studies. However, this retrospective design cannot establish causality, and residual confounding may persist despite matching and sensitivity analyses. Additionally, TRT-exposed patients may differ from unexposed patients in baseline health, endocrine evaluation, treatment eligibility, access to care, surveillance intensity, and comorbidity burden.

Additional limitations include incomplete capture of TRT dose, route, duration, adherence, indication, and achieved testosterone levels. KPS and ECOG were unavailable for many patients and could not be incorporated into our matching analyses. Serum total testosterone, free testosterone, sex hormone-binding globulin, estradiol, and glucocorticoid measurements were not available for large proportion of patients, limiting analysis of endogenous androgen biology or biochemical response to TRT. Corticosteroid dose, duration, and timing were also incompletely captured despite their relevance to the proposed HPA axis mechanism.

In summary, our findings demonstrate the association of TRT exposure with improved survival in male patients with glioblastoma and solid tumor CNS metastases. These findings are hypothesis-generating and should not be used to recommend TRT as an anti-tumor therapy outside standard endocrine indications. Prospective studies should incorporate standardized TRT (timing, dose, route, and duration), serial hormone measurements, corticosteroid exposure, performance status, tumor-specific stratification, and immune correlates to determine whether androgen signaling has a causal role in patients with primary and metastatic CNS tumors.

## Data Availability

All data produced in the present work are contained in the manuscript

## Required Statements

### Data Availability

Data will be made available upon request.

### Author Contributions

All authors contributed to the conception and intellectual framework of the manuscript, interpretation of the literature, drafting or critical revision of the manuscript, and approval of the final version. MMB performed all data analysis.

## References

1. Bernardez B, Higuera O, Martinez-Callejo V, et al. Sex and gender differences in cancer pathogenesis and pharmacology. Clin Transl Oncol. 2025;27(10):3837–3848.

2. Wang Z, Hu H, Bao Y, Ren G, Yang C. Sexual dimorphism in cancer: molecular mechanisms and precision oncology perspectives. Biol Sex Differ. 2026;17(1). doi:10.1186/s13293-026-00843-7

3. Lopes-Ramos CM, Burkholz R, Ben Guebila M, et al. Sex differences in gene regulation and its impact on cancer incidence. iScience. 2026;29(5):115570.

4. Vera R, Juan-Vidal O, Safont-Aguilera MJ, de la Peña FA, del Alba AG. Sex differences in the diagnosis, treatment and prognosis of cancer: the rationale for an individualised approach. Clin Transl Oncol. 2023;25(7):2069–2076.

5. Selvaraj RC, Cioffi G, Waite KA, Jackson SS, Barnholtz-Sloan JS. A pan-cancer analysis of age and sex differences in cancer incidence and survival in the United States, 2001–2020. Cancers (Basel). 2025;17(3):378.

6. Rubin JB, Lagas JS, Broestl L, et al. Sex differences in cancer mechanisms. Biol Sex Differ. 2020;11(1):17.

7. Tricarico R, Nicolas E, Hall MJ, Golemis EA. X- and Y-linked chromatin-modifying genes as regulators of sex-specific cancer incidence and prognosis. Clin Cancer Res. 2020;26(21):5567–5578.

8. Bustillos CG, Peluso EM, Cha SL, Lechner MG, Su MA. Sex matters: Hormonal and chromosomal determinants of autoimmunity and anti-cancer immunity across the lifespan. Immunol Rev. 2026;338(1):e70096.

9. Broestl L, Rubin JB. Sexual differentiation specifies cellular responses to DNA damage. Endocrinology. 2021;162(11):bqab192.

10. Ma J, Yao Y, Tian Y, Chen K, Liu B. Advances in sex disparities for cancer immunotherapy: unveiling the dilemma of Yin and Yang. Biol Sex Differ. 2022;13(1):58.

11. Dai S, Mo Y, Wang Y, et al. Chronic stress promotes cancer development. Front Oncol. 2020;10:1492.

12. Colon-Echevarria CB, Lamboy-Caraballo R, Aquino-Acevedo AN, Armaiz-Pena GN. Neuroendocrine regulation of tumor-associated immune cells. Front Oncol. 2019;9:1077.

13. Yu S, Gan C, Li W, et al. Depression decreases immunity and PD-L1 inhibitor efficacy via the hypothalamic-pituitary-adrenal (HPA) axis in triple-negative breast cancer. Biochim Biophys Acta Mol Basis Dis. 2025;1871(2):167581.

14. Curry RN, Glasgow SM. The role of neurodevelopmental pathways in brain tumors. Front Cell Dev Biol. 2021;9(659055). doi:10.3389/fcell.2021.659055

15. Price M, Ballard CAP, Benedetti JR, Kruchko C, Barnholtz-Sloan JS, Ostrom QT. CBTRUS statistical report: Primary brain and other central nervous system tumors diagnosed in the United States in 2018-2022. Neuro Oncol. 2025;27(Supplement_4):iv1–iv66.

16. Carrano A, Juarez JJ, Incontri D, Ibarra A, Guerrero Cazares H. Sex-specific differences in glioblastoma. Cells. 2021;10(7):1783.

17. Jovanovich N, Habib A, Chilukuri A, et al. Sex-specific molecular differences in glioblastoma: assessing the clinical significance of genetic variants. Front Oncol. 2023;13:1340386.

18. Bayik D, Zhou Y, Park C, et al. Myeloid-derived suppressor cell subsets drive glioblastoma growth in a sex-specific manner. Cancer Discov. 2020;10(8):1210–1225.

19. Lee J, Nicosia M, Hong ES, et al. Sex-biased T-cell exhaustion drives differential immune responses in glioblastoma. Cancer Discov. 2023;13(9):2090–2105.

20. Volovetz J, Alvarez-Vazquez A, Lee J, et al. Females experience a greater benefit from surgical resection in an immune-dependent manner in a preclinical glioblastoma model. Neurooncol Adv. 2026;8(1):vdag117.

21. Sloan AR, Bukenya G, Tannish GP, et al. Platelets regulate glioblastoma growth and immunity via sex-dependent PAR4 - Estrogen receptor beta signaling. bioRxivorg. Published online April 1, 2026. doi:10.1101/2025.08.06.668464

22. Tharp ME, Han CZ, Talukdar M, et al. The inactive X chromosome drives sex differences in microglial inflammatory activity in human glioblastoma. bioRxivorg. Published online May 19, 2025. doi:10.1101/2024.06.06.597433

23. Ochocka N, Segit P, Wojnicki K, et al. Specialized functions and sexual dimorphism explain the functional diversity of the myeloid populations during glioma progression. Cell Rep. 2023;42(1):111971.

24. Turaga SM, Silver DJ, Bayik D, et al. JAM-A functions as a female microglial tumor suppressor in glioblastoma. Neuro Oncol. 2020;22(11):1591–1601.

25. Lindenmaier Z, Yee Y, Kinman A, et al. Characterization of mice bearing humanized androgen receptor genes (h/mAr) varying in polymorphism length. Neuroimage. 2021;226(117594):117594.

26. Cara AL, Henson EL, Beekly BG, Elias CF. Distribution of androgen receptor mRNA in the prepubertal male and female mouse brain. J Neuroendocrinol. 2021;33(12):e13063.

27. Nguyen TV, McCracken J, Ducharme S, et al. Testosterone-related cortical maturation across childhood and adolescence. Cereb Cortex. 2013;23(6):1424–1432.

28. Anderson LC, Bolling DZ, Schelinski S, Coffman MC, Pelphrey KA, Kaiser MD. Sex differences in the development of brain mechanisms for processing biological motion. Neuroimage. 2013;83:751–760.

29. Wartenberg P, Farkas I, Csillag V, Colledge WH, Hrabovszky E, Boehm U. Sexually dimorphic neurosteroid synthesis regulates neuronal activity in the Murine brain. J Neurosci. 2021;41(44):9177–9191.

30. de Bournonville C, Lemoine P, Foidart JM, Arnal JF, Lenfant F, Cornil CA. Role of membrane estrogen receptor alpha (ERα) in the rapid regulation of male sexual behavior. J Neuroendocrinol. 2023;35(10):e13341.

31. Guan X, Polesso F, Wang C, et al. Androgen receptor activity in T cells limits checkpoint blockade efficacy. Nature. 2022;606(7915):791–796.

32. Consiglio CR, Udartseva O, Ramsey KD, Bush C, Gollnick SO. Enzalutamide, an androgen receptor antagonist, enhances myeloid cell-mediated immune suppression and tumor progression. Cancer Immunol Res. 2020;8(9):1215–1227.

33. Rodríguez-Lozano DC, Piña-Medina AG, Hansberg-Pastor V, Bello-Alvarez C, Camacho-Arroyo I. Testosterone promotes glioblastoma cell proliferation, migration, and invasion through androgen receptor activation. Front Endocrinol (Lausanne). 2019;10:16.

34. Alemán OR, Quintero JC, Hernández-Lúa LN, Espejel-Nuñez A, Camacho-Arroyo I. Testosterone-TGF-β crosstalk modulates cell migration in human glioblastoma-derived cell lines. Steroids. 2026;231–232(109798):109798.

35. Zalcman N, Canello T, Ovadia H, et al. Androgen receptor: a potential therapeutic target for glioblastoma. Oncotarget. 2018;9(28):19980–19993.

36. Guo G, Gong K, Wohlfeld B, Hatanpaa KJ, Zhao D, Habib AA. Ligand-independent EGFR signaling. Cancer Res. 2015;75(17):3436–3441.

37. Zalcman N, Larush L, Ovadia H, Charbit H, Magdassi S, Lavon I. Intracranial assessment of androgen receptor antagonists in mice bearing human glioblastoma implants. Int J Mol Sci. 2023;25(1):332.

38. Orevi M, Shamni O, Zalcman N, et al. [18F]-FDHT PET/CT as a tool for imaging androgen receptor expression in high-grade glioma. Neurooncol Adv. 2021;3(1):vdab019.

39. Lee J, Chung YM, Silver DJ, et al. Androgen loss accelerates brain tumour growth via HPA axis activation. Nature. 2026;653(8116):1184–1195.

40. Homepage. Mayo Clinic Platform. November 20, 2025. Accessed June 10, 2026. https://www.mayoclinicplatform.org/

41. Yu Y, Hu X, Rajaganapathy S, et al. Accelerating AI innovation in healthcare: real-world clinical research applications on the Mayo Clinic Platform. Npj Health Syst. 2026;3(1):17.

42. Foskolou IP, Bunse L, Van den Bossche J. 2-hydroxyglutarate rides the cancer-immunity cycle. Curr Opin Biotechnol. 2023;83(102976):102976.

43. Ježek P. 2-hydroxyglutarate in cancer cells. Antioxid Redox Signal. 2020;33(13):903–926.

44. Bader DA, Chakraborty B, McDonnell DP, Hirschey MD. Targeting androgen receptor signaling to enhance cancer immunotherapy. Trends Pharmacol Sci. Published online December 3, 2025:S0165-S6147(25)00257–3.

45. Thon N, Kreth S, Kreth FW. Personalized treatment strategies in glioblastoma: MGMT promoter methylation status. Onco Targets Ther. Published online September 2013:1363.

46. Poon MTC, Keni S, Vimalan V, et al. Extent of MGMT promoter methylation modifies the effect of temozolomide on overall survival in patients with glioblastoma: a regional cohort study. Neurooncol Adv. 2021;3(1):vdab171.

47. Pettersson-Segerlind J, Mathiesen T, Elmi-Terander A, et al. The risk of developing a meningioma during and after pregnancy. Sci Rep. 2021;11(1):9153.

48. Szulzewsky F, Thirimanne HN, Holland EC. Meningioma: current updates on genetics, classification, and mouse modeling. Ups J Med Sci. 2024;129. doi:10.48101/ujms.v129.10579

49. Berghaus N, Hielscher T, Savran D, et al. Meningiomas: Sex-specific differences and prognostic implications of a chromosome X loss. Neuro Oncol. 2025;27(4):1019–1028.

50. Park JS, Sade B, Oya S, Kim CG, Lee JH. The influence of age on the histological grading of meningiomas. Neurosurg Rev. 2014;37(3):425–429; discussion 429.

51. Buerki RA, Horbinski CM, Kruser T, Horowitz PM, James CD, Lukas RV. An overview of meningiomas. Future Oncol. 2018;14(21):2161–2177.

52. Felistia Y, Amanda NF, Hendrawan F, Susanto NH, Al Fauzi A, Miftahussurur M. Retrospective analysis of recurrence patterns and clinical outcomes in grade I-III meningiomas after surgery. Surg Neurol Int. 2025;16:149.

53. Durand A, Labrousse F, Jouvet A, et al. WHO grade II and III meningiomas: a study of prognostic factors. J Neurooncol. 2009;95(3):367–375.

