## Supplemental Table 1 for "Protective effects of testosterone replacement therapy on brain tumor outcomes: the Mayo Clinic Experience"

| Table 1. Baseline Characteristics by Tumor Cohort and TRT Exposure | | | |
|---|---|---|---|
| **Variable** | **Brain metastases - No TRT** | **Brain metastases - TRT** | **GBM - No TRT** |
| N | 5237 | 91 | 587 |
| Age at diagnosis, median (IQR) | 66.0 (57.9-73.1) | 67.7 (61.2-74.1) | 64.6 (56.7-70.8) |
| Survival time, months, median (IQR) | 7.1 (2.4-20.9) | 14.1 (5.0-36.1) | 13.2 (7.9-20.8) |
| Deaths | 4250 (81.2%) | 71 (78.0%) | 510 (86.9%) |
| Male | 5237 (100.0%) | 91 (100.0%) | 587 (100.0%) |
| Hispanic or Latino | 4112 (78.5%) | 75 (82.4%) | 569 (96.9%) |
| Not Hispanic or Latino | 3999 (76.4%) | 73 (80.2%) | 541 (92.2%) |
| Unknown/Other ethnicity | 1123 (21.4%) | 16 (17.6%) | 16 (2.7%) |
| MGMT methylated | 0 (0.0%) | 0 (0.0%) | 198 (33.7%) |
| MGMT unmethylated | 0 (0.0%) | 0 (0.0%) | 389 (66.3%) |
| Primary cancer: Lung | 3001 (57.3%) | 43 (47.3%) | 0 (0.0%) |
| Primary cancer: Melanoma | 955 (18.2%) | 21 (23.1%) | 0 (0.0%) |
| Primary cancer: Renal | 538 (10.3%) | 17 (18.7%) | 0 (0.0%) |
| Primary cancer: Colorectal | 283 (5.4%) | 4 (4.4%) | 0 (0.0%) |
| Primary cancer: GI/Hepatobiliary/Pancreatic | 275 (5.3%) | 2 (2.2%) | 0 (0.0%) |
| Primary cancer: Urothelial | 103 (2.0%) | 2 (2.2%) | 0 (0.0%) |

| | | | |
|---|---|---|---|
| Primary cancer: Sarcoma | 82 (1.6%) | 2 (2.2%) | 0 (0.0%) |
| KPS 80-100 | 508 (9.7%) | 11 (12.1%) | 210 (35.8%) |
| KPS 60-70 | 122 (2.3%) | 2 (2.2%) | 52 (8.9%) |
| KPS <60 | 35 (0.7%) | 0 (0.0%) | 17 (2.9%) |
| KPS unknown/missing | 4572 (87.3%) | 78 (85.7%) | 308 (52.5%) |
| ECOG 0-1 | 2273 (43.4%) | 47 (51.6%) | 334 (56.9%) |
| ECOG 2 | 480 (9.2%) | 9 (9.9%) | 53 (9.0%) |
| ECOG 3-5 | 307 (5.9%) | 2 (2.2%) | 31 (5.3%) |
| ECOG unknown/missing | 2177 (41.6%) | 33 (36.3%) | 169 (28.8%) |
| Extracranial metastatic disease | 2652 (50.6%) | 56 (61.5%) | 0 (0.0%) |
| Radiation procedure/exposure | 3594 (68.6%) | 63 (69.2%) | 409 (69.7%) |
| SRS | 1894 (36.2%) | 36 (39.6%) | 0 (0.0%) |
| Surgery | 764 (14.6%) | 16 (17.6%) | 504 (85.9%) |
| Local therapy | 3800 (72.6%) | 65 (71.4%) | 409 (69.7%) |
| Temozolomide exposure | 0 (0.0%) | 0 (0.0%) | 464 (79.0%) |
| Dexamethasone exposure | 0 (0.0%) | 0 (0.0%) | 516 (87.9%) |
| Immune checkpoint inhibitor exposure | 1174 (22.4%) | 36 (39.6%) | 0 (0.0%) |

*Values are median (IQR) or n (%). KPS, ECOG, and ICI are shown when available.*

| |
|---|
| GBM - TRT |
| 11 |
| 68.2 (62.2-72.7) |
| 12.6 (9.1-20.4) |
| 7 (63.6%) |
| 11 (100.0%) |
| 2 (18.2%) |
| 9 (81.8%) |
| 0 (0.0%) |
| 4 (36.4%) |
| 7 (63.6%) |
| 0 (0.0%) |
| 0 (0.0%) |
| 0 (0.0%) |
| 0 (0.0%) |
| 0 (0.0%) |
| 0 (0.0%) |

| |
|---|
| 0 (0.0%) |
| 6 (54.5%) |
| 0 (0.0%) |
| 1 (9.1%) |
| 4 (36.4%) |
| 6 (54.5%) |
| 2 (18.2%) |
| 0 (0.0%) |
| 3 (27.3%) |
| 0 (0.0%) |
| 9 (81.8%) |
| 0 (0.0%) |
| 10 (90.9%) |
| 9 (81.8%) |
| 8 (72.7%) |
| 8 (72.7%) |
| 0 (0.0%) |
